# Lived experiences and the effects of domestic violence on female secondary schoolteachers in Uganda: A qualitative study using in-depth interviews

**DOI:** 10.1101/2023.08.27.23294238

**Authors:** Gladys Ayot Oyat, David Lagoro Kitara, Mary Ocheng Kagoire, Wilson Eduan

## Abstract

**Background:** Domestic violence is a multifaceted and complex phenomenon that, in many ways, affects victims and their workplaces. Little is known about lived experiences and the effects of domestic violence on female secondary schoolteachers in Uganda. This study explored lived experiences and the effects of domestic violence on female secondary schoolteachers on their professional and administrative roles.

**Methods:** Underpinned by a feminist paradigm, the study employed a qualitative research approach and narrative design. Snowballing was used to a point of theoretical saturation. In-depth interviews were conducted with twenty female secondary schoolteachers who were victims of domestic violence perpetrated by their spouses and triangulated with interviews of twenty-three headteachers from schools in northern Uganda. Conceptualization and meanings were derived from experiences of female teachers, which contributed to the understanding of complex issues of domestic violence and how it affects their professional and administrative roles. A local IRB approved this study, and narrative, thematic analysis was used.

**Results:** The finding indicates that female secondary school teachers have lived experiences of domestic violence, affecting their professional and administrative roles, resulting in poor service delivery and interpersonal relationships. The effects were poor service delivery, absenteeism, missing lessons, poor lesson preparations, low self-esteem, mental health issues, depression, poor interpersonal relationships, bad attitudes, and lack of cooperation with others. Female teachers used both formal and informal means to cope with the situation.

**Conclusion:** This exploratory study brings awareness to the lived experiences and effects of domestic violence on female secondary schoolteachers in Uganda. Domestic violence affects their professional and administrative roles with resulting poor service delivery and poor interpersonal relationships. Providing a supportive work environment is crucial for female teachers experiencing domestic violence to enable them to perform their professional and administrative roles. Different players, school managers, the Ministry of Education and Sports, feminist organizations, and relevant government offices can intervene by establishing workplace policies and relevant processes to support female teachers who experience domestic violence.

## Background

Domestic violence is a multifaceted and complex phenomenon that affects the victims and their workplaces in many ways.^1^ It is a widespread global issue that crosses boundaries such as socioeconomic class, race, region, and religion.^2^ Globally, it has become an essential field of research in the last decade and has emerged as a global trend promoted by the United Nations, Human Rights activists, and feminist movements.^3^ The World Health Organization (WHO) has pronounced violence against women an endemic phenomenon.^3^ Based on regional variance, its prevalence ranges from 45% among women in the African continent to 27.2% in Europe, and about 30% of women get affected by domestic violence at some point in their lives.^3^

Rayner-Thomas et al., 2013 found that 13 to 61% of women in ten countries experienced some form of physical violence from a male partner, 6 to 59% experienced sexual violence, and 20-75% experienced emotional and psychological violence.^4^

According to statistics from the Malaysian Royal Police to Parliament, 62,670 domestic violence cases were reported from 2000 to 2017, with an estimated number of 3800 cases per year, equivalent to 323 cases per month.^5^ Additionally, the National Crime Record Bureau in 2011 reported 8,618 dowry deaths in India, but unofficial figures estimated that dowry deaths were three times more in India than previously thought.^6^

Domestic violence is a shared experience among the working class. In a study conducted on domestic violence among white-collar working women in Turkey, 10,000 white-collar working women were found to have experienced severe physical violence from their partners every year.^7^ This finding perhaps confirms that domestic violence affects all women irrespective of their status, religion, position, or economic level, with no exceptions to female teachers.

In sub-Saharan Africa, empirical evidence on the prevalence of domestic violence is limited and confined to a few population studies.^8^ In 1995, Egypt Demographic and Health Survey (EDHS) showed that 35% of women were beaten by their husbands.^9^

In Uganda, many efforts have been invested in ending domestic violence against women in communities. It developed a solid normative legal framework on gender-based violence and its harmful practices.^10^ For example, Uganda’s 1995 Constitution plus a broader normative and policy frameworks reflect global standards and strongly support gender equity in communities.^10^ The National Action Plan on Elimination of Gender-based Violence in Uganda (2016-2020) frames the issue of gender-based violence as an urgent development priority and a factor to be addressed in achieving Uganda’s development goals.^10^

Although gender issues in Uganda lean more toward females and issues of domestic violence are fully catered for in the law, many laws do not address critical aspects of violence against women.^10^

The Uganda Domestic Violence Act No. 17 of 2010 puts in place stringent protection measures for domestic violence victims.^11^ It defines domestic violence broadly to include physical, sexual, verbal, psychological, emotional, and economic abuse of a victim or anyone related to him or her in a household.^11,12^

In addition, the national policy on the elimination of Gender Based Violence (GBV) in Uganda and the associated National Plan of Action on elimination of Gender Based Violence in Uganda (2016) are aligned with all relevant policies and help summarise the broad body of national laws, policies, and international commitments which address Gender Based Violence.^13^ These policies focus most attention on ending impunity for perpetrators of violence and creating an enabling environment for accountability of stakeholders.^13^ However, the prevalence of domestic violence in many communities in Uganda is still high.^11,12,13^

Reports from Non-Governmental Organizations (NGOs), media, and police show that women suffer more from domestic violence than men.^14^ In a 2009 study by the Uganda Law Reform Commission (ULRC), one-half of women reported experiencing violence daily or weekly, and 35% of working women reported marital violence.^14^

Furthermore, the Uganda Demographic Household Survey (UDHS, 2011)^15^ and Uganda Bureau of Statistics (UBOS, 2011)^16^ indicate a high prevalence of gender-based violence with an overall prevalence rate by type of violence at 56% for physical violence, 27.7% for sexual violence, and 42.9% for spousal violence.^15,16^ These figures show a slight improvement from the Uganda Demographic Health Survey (UDHS, 2016)^15^, which indicates that a portion of ever-partnered women aged 15-49 experienced intimate partner physical or sexual violence in a lifetime at 50% and that 39.6% had experienced it within the last one year.^17^

In the district of study, several reports on the civil conflict in Northern Uganda have noted domestic violence as one of the most persistent violations of women’s and girls’ rights and a significant public health problem in the region.^18^ The lived experiences of domestic violence among female secondary schoolteachers and how it affects their professional and administrative roles remain to be confirmed and brought to light.

Black et al. (2019) asserts that the prevalence of domestic violence is high in Northern Uganda, with high rates of emotional, physical, and sexual violence, including 78.5% of women having experienced at least one type of domestic violence, and more than half having experienced domestic violence in the twelve months prior to their survey.^14^

This study explored lived experiences and the effects of domestic violence among female secondary schoolteachers on their professional and administrative roles.

## Methods

### Study design

This was a cross-sectional study conducted using qualitative research methods.

### Study Settings

This study was part of a larger project demonstrating lived experiences of domestic violence among female secondary schoolteachers in Uganda. It focused on assessing female secondary schoolteachers’ perspectives on how domestic violence affects their professional and administrative roles. An in-depth interview method was used to address the research questions and a narrative research design was employed to explore experiences of female secondary schoolteachers to describe the meaning of individuals’ experiences^19^ and obtain multiple perspectives that will add more understandings on domestic violence and how it affects female teachers’ professional and administrative roles.

### Participants

Female secondary schoolteachers with lived experiences of domestic violence were the target of our research, beginning with the ones already known by the researcher and were willing to be interviewed. The team interviewed twenty female secondary schoolteachers with experiences of domestic violence, past and ongoing, and working in secondary schools in the Kitgum district of northern Uganda. In addition, twenty-three secondary school headteachers in the same district were interviewed to triangulate the information obtained from the female teachers.

### Recruitment and interviews of participants

A progressive recruitment of participants was conducted by snowballing method to identify participants until a point of theoretical saturation was reached. In this, the scope of the study was limited to female secondary schoolteachers in the district, who were victims of domestic violence and headteachers from their respective secondary schools.

The research team employed in-depth interviews where data was collected using face-to-face questionnaire interviews. The triangulation of information was achieved by interviewing headteachers of their respective secondary schools. The researcher used an open-handed interview guide to direct the conversation and allow participants to provide information to help them understand the in-depth and lived experiences of domestic violence to themselves and its effects on their professional and administrative roles.

### Ethical approval

This study was approved by the Research and Ethics Committee (REC) of Uganda Christian University (UCU) and Uganda National Council for Science and Technology (UNCS&T). In addition, the Kitgum District Education office authorized the conduct of this study, where all participants provided written informed consent to participate. In addition to participants’ written informed consent, confidentiality, respect for participants’ privacy, and keeping personal information confidential were ensured. Although the face-to-face interviews were conducted with participants during the peak of the COVID-19 pandemic when the risk of getting infected with COVID-19 could have been high due to close physical interactions, we ensured that we adhered to the national infection, prevention, and control (IPC) and Standing Operating Procedures (SOPS) for COVID-19 by wearing facemasks, keeping social distancing, washing hands, and sanitizing before, during and after interviews.

### Data analysis

The primary data obtained for this study was recorded (written and audio recorded) and analyzed using thematic narrative analysis.^19,20^ Thematic experience-centered narrative analysis was used to identify themes and sub-themes within the narratives.^19,20^ The researcher analyzed data manually because of the number of participants and the information obtained. The author became familiar with the data by reading and re-reading all written materials and listening repeatedly to audio recordings from the interviews.^19,20^ The researcher then generated initial codes, searched for themes, reviewed them, and later defined and named the theme before reporting.^19,20^ Inductive coding was created based on the data and labels were created as they emerged.^19,20^ Consequently, the analysis on lived experiences and effects of domestic violence on female secondary schoolteachers’ professional and administrative roles was maintained inductive throughout the analysis process.

### Credibility

To achieve a high standard of credibility, honesty, trustworthiness, and the purpose of this study was made known to participants. A suitable venue was agreed upon with participants’ consent before the study’s commencement. The credibility of the study’s findings was ensured by selecting participants who had experienced domestic violence, the use of in-depth interview techniques for data collection, and the analysis of findings using narrative design. In addition, we established a data trail, acknowledging the researcher’s subjectivity, conducting participant checks and reviews, and ensuring prolonged engagements and follow-up with participants on the subject matter.

### Results of our findings

The study findings were on lived experiences of domestic violence among female secondary schoolteachers in Northern Uganda. It described how domestic violence affected their professional and administrative roles in the in-depth interviews. The interviewed participants’ sociodemographic characteristics were ages, qualifications, number of children, subjects taught, and years in marriage (Table 1). In addition, similar information was recorded for headteachers in secondary school in Kitgum including their experience as headteachers (Table 2).

**Table 1:**
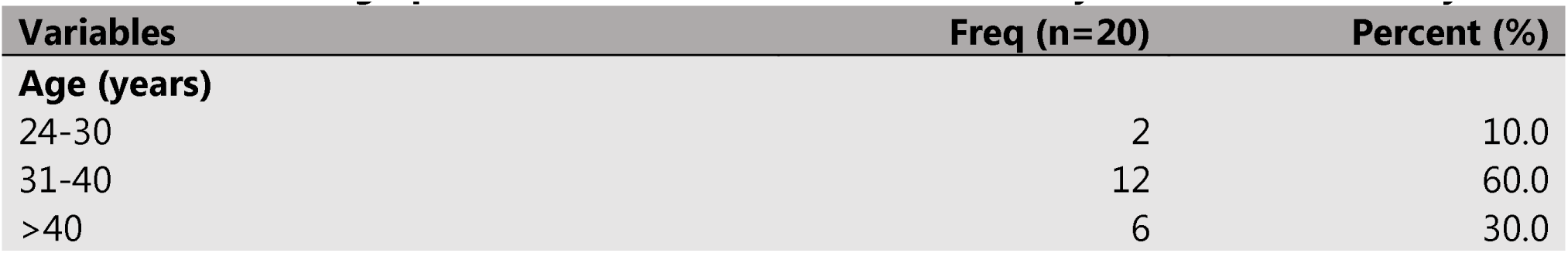
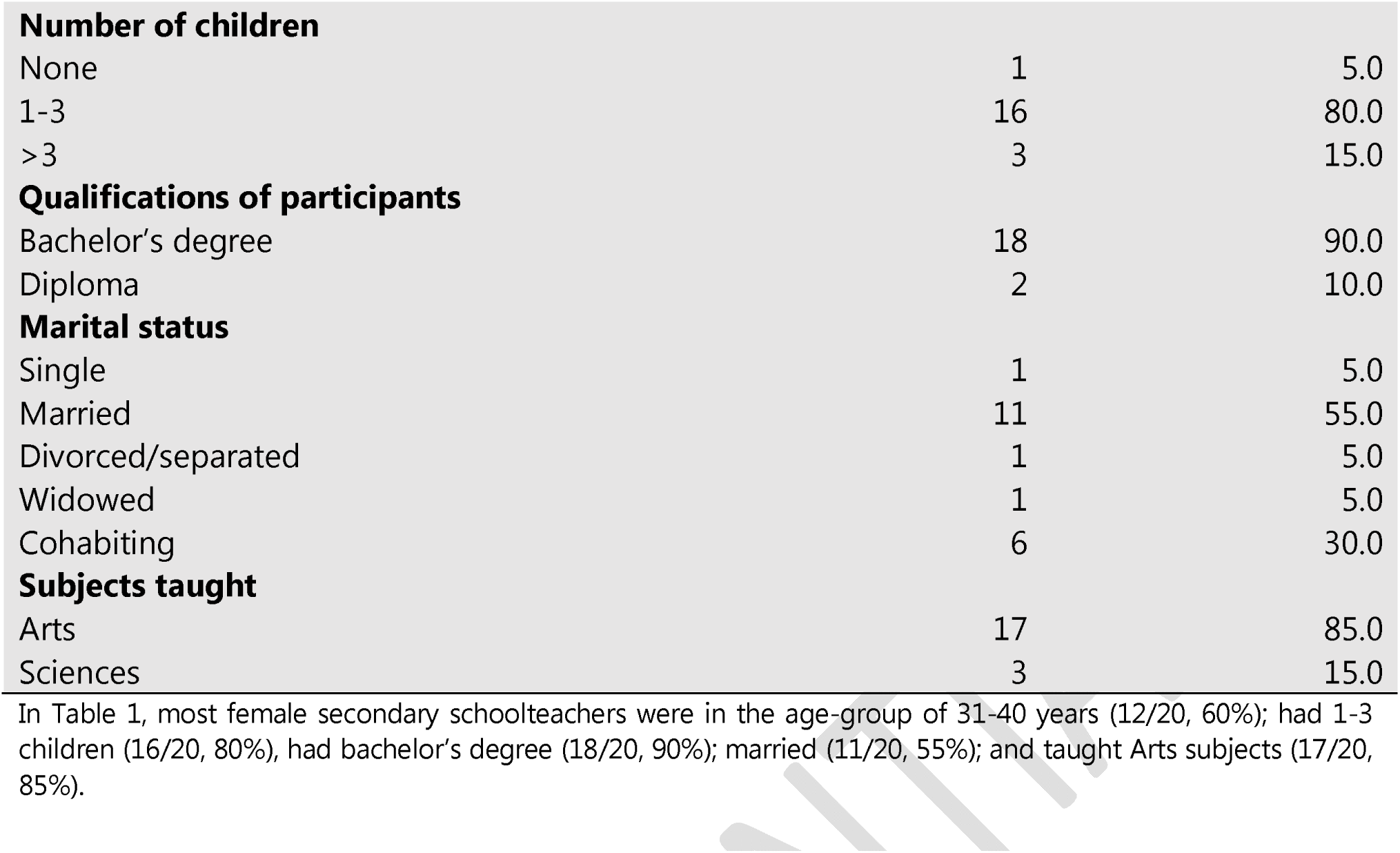
Sociodemographic characteristics of female secondary teachers in our study.

**Table 2:**
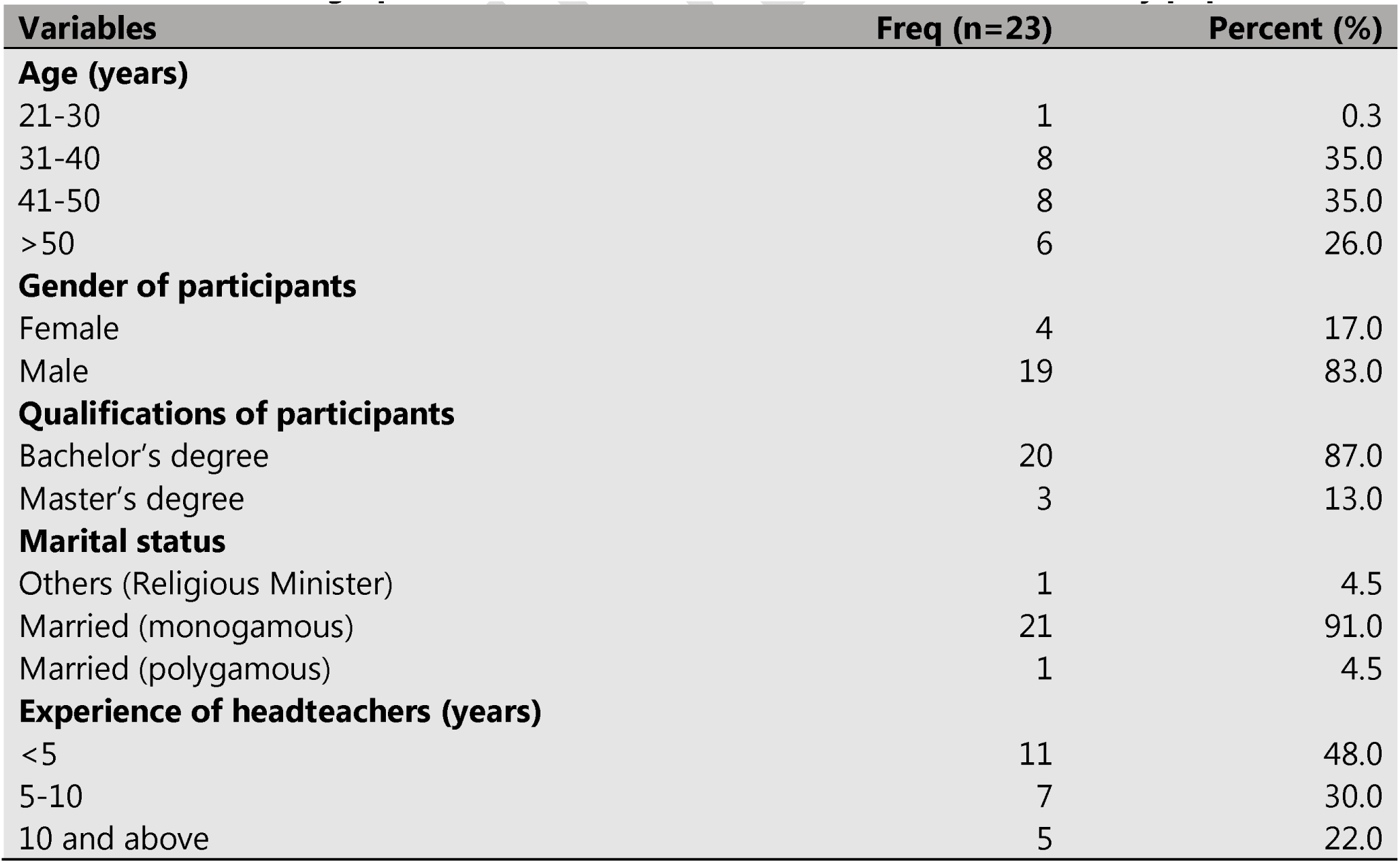
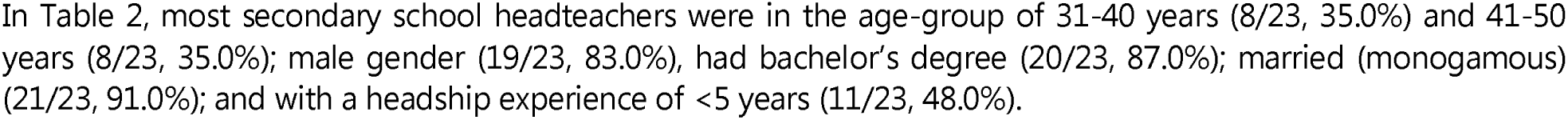
Socio-demographic characteristics of the headteachers in the study population.

**Table 3:** The themes and sub-themes for this research question.

|  |  |
| --- | --- |
| **Theme 1: Poor service delivery.** | ▪ **Sub-themes**<br>✓ ***Absenteeism***<br>✓ Missing lessons<br>✓ Poor lesson preparation<br>✓ Low self -esteem<br>✓ ***Mental health problems*** |
| **Theme 2: Poor interpersonal Relationship.** | ▪ **Sub-themes**<br>✓ Negative attitudes<br>✓ Lack of cooperation |

### Background information on female secondary schoolteachers

Twenty participants in this study had or were experiencing domestic violence and were willing to discuss their lived experiences (Table 1). Accordingly, (2/20, 10.0%) female teachers were aged 25-30 years. Most participants (12/20, 60.0%) were aged 31-40 years, comprising 60% of the participants. Women aged 40 and above were (6/20, 30.0%) (Table 1). Reviewing this age group distribution, it can be argued that between the ages of 25-30 years, female teachers were less willing to talk about their family challenges. This age group is probably tender with many family and relation expectations. Therefore, at this age group, spouses try to please each other, and the relationship leaves room for tolerance and violence.

According to the Ugandan education system, most students complete their university or tertiary education between the ages of 20 and 24 years. After that, they take a few years before getting married, probably accounting for the lower percentages of domestic violence than in older age-groups in this study population. In addition, lower chances and loopholes presented by snowballing technique may be a reason for lower percentages of domestic violence in this age group than the others.

The age group of 31 to 40 years had the highest percentage of domestic violence in this study population (Table 1). The possible explanation could be that women in this age group were more confident and willing to talk freely about the pain they suffer in their families without consideration on the impact on their families if their spouses got to know (Table 1). The percentage drop in prevalence in the age group above 40 years was probably due to female teachers being more independent and assertive. Besides, being married for many years may have bred some level of tolerance and coping with the situation. Middle-aged couples were either more prone to domestic violence or were willing to report violence inflicted on them (Table 1).

Further, most female teachers who suffered domestic violence had one to three children, constituting (16/20, 80.0%). Those with more than three children were (3/20, 15.0%), and one (1/20, 5.0%) had no child in the marriage (Table 1). Most participants (11/20, 55.0%) were married; (1/20, 5.0%) separated; (1/20, 5.0%) divorced; (1/20, 5.0%) widowed, and (6/20, 30.0%) cohabiting. Aggregating these numbers, (16/20, 85.0%) of the teachers continued to live in abusive relationships without divorcing their husbands. The main reasons for these were “staying for the sake of children,” “cultural stigma,” and the “fear of the unknown.”

On the other hand, a female teacher in her 50s, who was no longer living in abusive relationship said she saw no reason for staying childless in a dysfunctional relationship. This finding confirms the importance of children for the perseverance of female teachers even in a dysfunctional relationship afflicted by violence.

Most female teachers reported having attained a bachelor’s degree and comprised (18/20, 90.0%) of the female teachers in our study population. The level of education did not change the experience of domestic violence in their families.

Also, we found that domestic violence occurred among female secondary schoolteachers across all age groups, marital statuses, and educational levels. Children in marriage played a crucial role in convincing women to persevere in their violent marital relationships (Table 1).

### Background information on secondary school headteachers

Twenty-one (21/23, 91.0%) headteachers were in a monogamous marriage; (1/23, 4.5%) in a polygamous, and (1/23, 4.5%) was a religious Minister. Eight (8/23, 34.7%) were in the age group of 31 to 40 years, and the same number (8/23, 34.7%) in 40 to 50 years. Six (6/23, 26%.0) were above 50 years, and one (1/23, 4.3%) was below 30 years (Table 2). Of these, only four (4/23,17.3%) were females, and (19/23, 82.6%) were males. It was expected that more profound knowledge and support for the victims of domestic violence from female headteachers. However, we were surprised that fewer female secondary schoolteachers confide in their female headteachers than their male counterparts. The reason was that female headteachers were less trusted and not closely related to their staff (Table 2). Some participants said female headteachers were not approachable and therefore it did not serve any purpose discussing their sensitive challenges. Female schoolteachers aged 45 years and above were more likely to share their domestic violence experiences with their female headteachers. One gave the reason that female headteachers were supportive, like an elder sister or a mother. However, a few younger female schoolteachers believed that female headteachers would be of help, except one (Table 2). Most secondary school headteachers (11/23, 47.8%) had the experience of being headteachers for less than five years. Most headteachers with less experience were less likely to show any reason for supporting female teachers in domestic violence relationships. In addition, most headteachers agreed that they knew domestic violence was among staff, although they had not received it directly from the female teachers.

According to headteachers, several female teachers were employed in their schools, the highest number in a school being thirteen and the lowest, one. At the time of the study, ninety-eight female secondary school teachers reported to be working in the twenty-three secondary schools in Kitgum district. They knew that many female teachers were experiencing domestic violence in their families (Table 2).

### Origins of domestic violence as reported by female secondary schoolteachers

Female teachers were asked what makes them happy in their marriage. There were several themes that emerged from the discussions. These included good communication with their partners, material support, being welcomed by in-laws, having a hardworking partner, having a God-fearing partner, a caring partner, a partner who provides for the family, and siring children. Even where domestic violence was evident, some factors made female teachers appreciate their abusive spouses, thereby persevering in the marriage.

### Several issues were pointed out regarding the reasons for domestic violence in their relationship

These included extramarital affairs (15/20, 75.0%); lack of financial support/financial exploitation (20/20, 100.0%); not being trusted (18/20, 80.0%), physical fights/power control (15/20, 75.0%); not being allowed to work, being hot-tempered; adverse influence from husband’s family members; differences in educational level; stalking and emotional abuses such as sexual deprivation; disrespectful verbal utterances, and disrespectful actions.

Headteachers of secondary schools confirmed that these reasons presented by their female teachers were true. Four (4/23, 17.3%) confirmed that they were aware that female teachers in their schools experienced physical abuse mainly arising from husbands wanting to be in control/power; seven (7/23, 30.4%) said that the female teachers reported emotional, psychological abuse, eight (8/23, 34.8%) reported economic exploitation, and another (1/23, 4.3%) extramarital affairs.

### Issues of culture, level of education, Christian faith, and extramarital affairs

In the study, most women explained that domestic violence in their relationships had its root causes in extramarital affairs. The majority said they always felt unloved, inadequate, jealous, angry, and insecure when they find out their husbands were in other relationships. These extra-marital affairs affect their self-esteem because of the feeling of deep betrayal, disrespect, and failure to recognize their contributions to the marriage.

Being educated Christians, they feel that polygamy is backward; that it is a tradition that should not apply to women of their status, and that such awful culture should be eradicated. They said it would result in family tensions whenever they oppose these actions and men insist on their way.

The violence usually results from the “cold” response to their husbands, who expect them to remain happy and welcoming even when they were aware of what was happening (extramarital affairs). They reported provoking their husbands out of jealousy, as nothing remains the same when a spouse is in another relationship. Jealousy, hate, insecurity, mistrust, and change of attitudes by the affected female teachers contribute to the tensions, misunderstandings and resulting violence. For example, extramarital affairs as the reason for domestic violence was given by FV. 20, a female teacher in her 30s, said.

> *Madam, as I said, when my husband wants to have sex when I know he has been sleeping with another woman, I usually deny him. When that happens, he reacts by beating, abusing, and he stops providing for the household, yet he expects me to prepare food and wash his clothes.*

All (20/20,100.0%) female teachers and six (6/23, 26.1%) headteachers mentioned the lack of financial support and financial exploitation as sources of disagreement and violence in households. As reported by female teachers on domestic violence, financial issues were very diverse. Many men were unsupportive financially, most responsibilities were left to women, particularly for buying food, cultivating, paying utilities, meeting costs of children’s scholastic materials, and hospital bills. In extreme cases, female teachers paid all costs of school fees, housing, and demands from extended families. Such economic exploitations bred tensions in most female teachers, often leading to physical and emotional abuse.

Another source of conflict experienced by female teachers was lack of financial support. This finding points to situations when husbands pick money from their wives without consent, and in many cases leads to fights and violence. Women felt that failing to support them financially was already bad, but going further to take their money without permission was very painful and unacceptable. Besides, this action led to a feeling of lack of respect and privacy. Such monies were picked from handbags, bottles, containers for local household savings, access to bank ATM cards, and withdrawing money from their bank accounts. Quarrels and physical fights commonly followed such actions.

One participant, FV.15, lamented,

> *Even when he does not give me money, I have found him checking [checking in] my handbag more than three times and picking my money. When asked whether he picked [up] money from my handbag, he agreed and said: “Yes, I picked [up] the money that your boyfriend sent you”.*

Such statements point to the economic violence combined with verbal utterances that may psychologically affect female teachers. They increase the pain more if female teachers were not happy with what the money was used for. Most times, after picking the money, they were used for the wrong reasons, such as drinking alcohol, hanging out with friends, eating pork, and giving to girlfriends. To many, it would be understandable if the money was used within the family.

Another teacher, FV.13, in her 40s, had the same feelings and said,

> *“I was used as a ladder. I paid for his degree course. After finishing his course, he married another woman and now lives with her…. I paid rent for my mother-in-law for three years while he was in school, and I supported him and his family. All I received for doing good is pain and rejection… my pains cannot go away, and I can do nothing, but God will never bless them. By the way, even my mother-in-law who pretended all along to like me, told me to allow the son to be fully in charge of everything. When I said no, she also turned against me.”*

The issue of financial exploitation is known by the headteachers as well. Some considered this a significant challenge to women, and it was the reason for many spousal problems. Although many headteachers agreed that financial exploitation could cause domestic violence and that men should support their wives financially, the researcher also registered adverse voices from male headteachers who acted as though they were supporting domestic violence. Such headteachers think that women’s consistent fight for equality in Uganda should be followed with accountability so, there was nothing wrong if female schoolteachers’ husbands did not support them financially since they also earn salary.

A male headteacher in his 40s said, *“These women want to be equal. Why don’t they confirm this through their financial muscles?”*

> *Another headteacher said, “Let these women know very well that men are heads of the family and culturally no woman owns anything in a home and so when a man utilizes what the woman has brought home there must be no resistance, and no quarrels…”*

Such positions depict the patriarchal nature of our society, and it was doubtful that female teachers could get support from such headteachers to enable them to cope with domestic violence as they performed their professional and administrative roles. Listening to the female teachers’ stories, the cultural feelings that men own everything and women nothing came into play, and on the other side, feelings that men should be providers also remained strong in the narrative.

From female schoolteachers’ perspectives, another reason for domestic violence was the lack of trust. For peaceful and harmonious coexistence between spouses, trust must be at the core of the marriage. From the women’s stories, domestic violence arose when there was mistrust between spouses. Trust must be from both wife and husband. According to female teachers, suspicion always came from actions of their husbands. There were several reasons why such actions eroded the trust in the family.

All women whose spouses had lower academic standards than them reported a lack of trust by their husbands. They said their spouses think they were not faithful when they delay in schools, attend school functions like parties, walk with male teachers, or dress smartly. Their husbands usually felt insecure and jealous. Such lack of trust degenerated into emotional violence, abuse, restriction of movements, isolating, and often leading to physical fights.

Female teachers agreed that once their husbands began to lie or when there was evidence of unfaithfulness, they constantly dwelled on that, and this led to violence. Several participants reported having been uncomfortable with not being trusted by their husbands.

A young woman, FV.3, said,

> *He is a very jealous and not an enlightened man. He does not trust me at all… One day when I came from work, I found him in a horrible mood, and he told me that he saw me moving with men. He said you surround yourself with men like a dog on heat. Even a dog is better than you.*

She went on to state this amidst sobs,

> *When I tried to respond that those males were colleagues from the school, and we were working together to prepare students for athletic[s]. He reacted by slapping, kicking, and insulting me severely. I cried not because of the pain from the blows and kicks but from the verbal insults I am innocent, I have never cheated on him. Despite all my assurance he continued saying that I have sex with male teachers. That I sleep with students and men who advance on me.*

Such utterances from a spouse were bound to affect the performance of a female teacher. To them, even when the husbands were not around, thinking about such utterances caused pain and affected their preparation and delivery of lessons.

Generally, the lack of trust by husbands stemmed from allegations of unfaithfulness of their wives. They reported that their husbands were always suspicious and accused them of infidelity, which, to them, was not true. These accusations were mainly experienced by younger schoolteachers. Tapping mobile phone calls and checking spouses’ call records and messages were reported by female teachers in their 30s or below. These actions showing mistrust made female teachers to get agitated and led to violence. In addition, the lack of trust sparked emotional and physical abuse, eventually escalating to a feeling of financial exploitation and deprivation.

One female victim, FV14, in her late 40s, said,

> *The man is secretive. He feels that there are things I should not know. When asked, we always end up quarreling and physical fights. If I am his wife and have intimacy with him, why must he keep secrets from me, and how can I trust that I can be safe with him.*

While trust issues were common reasons for conflict among younger and older female teachers, the younger women struggled with the lack of trust, mainly from suspicion of unfaithfulness and bad friends. In contrast, women in their 40s and above had issues of trust mainly on resources and how they were managed.

### Power and control issues were reported as another cause of domestic violence

This study showed that (15/20, 75%) female teachers reported that they had been beaten, slapped, pinched, kicked, and boxed by their spouses simply because their husbands felt they must be in control. From the conversations, they were unhappy that at their level of education, their husbands could still beat them to the extent of sustaining physical injuries, which left them in pain, shame, and absent from work. Such fights were unexpected, and occurred when husbands felt their authority were being questioned. In addition, a single slap, or any physical fight, bred domestic violence because it bred anger because of the disrespect. Besides, most often female teachers responded violently in self-defense, which led to more cycles of violence in the household.

### Some of these fights had always left female teachers with physical injuries that resulted in hospitalization

Female teachers explained that the physical scars were embarrassing, and, in most cases, they had to look for reasons to hide the incident by saying they got hurt after falling. After sustaining fresh injuries from the violence, they usually stayed away from school, affecting their work, while their husbands felt powerful and in control.

Of the 23 headteachers, four (4/23, 17.4%) confirmed that female teachers suffer from physical violence. They agreed that the patriarchal feelings among husbands lead them to feel they must be in control, and women who challenge this position risks domestic violence. In contrast, the number of female teachers who agree with that position was far less. This finding is likely because female teachers who suffer from domestic violence hide this information from their supervisors because of the embarrassment and shame that the revelation can bring.

### Harmful Influence of family members was another factor leading to domestic violence

In the study area, family affairs are still guided by traditional African cultures. Women who are married in a home must pay allegiance to a broader family, and there was a lot of Influence from family members on how their spouses live. The family members include mothers-in-law, fathers-in-law, aunties, uncles, and siblings. These extended families including relatives were a complex situation and depended on the nature of the families where the women were married. Some family members did not want female teachers to work, while others felt they must not complain. They expected women to obey their husbands without complaints at all times. Women’s ideas were secondary and did not matter in family settings, while other relatives believed that married women should not be involved in family meetings because they were foreigners in family matters; they must not support her family because she had already been married. Others were extreme by directing female teachers to be good wives by cooking and serving all meals to their husbands and close relatives. Yet, it did not matter to them whether female teachers had to be in schools performing their work or not.

To female schoolteachers, such demands became overwhelming physically, mentally, and emotionally when they saw their husbands were not protecting them from relatives. They complained, and, in many cases, this led to violence, affecting their relationship and school workplaces. For example, in the name of being good wives, they tried to balance their work and home duties, but where there was no understanding, they ended up pleasing neither side. In contrast to many situations, some relatives became jealous and felt their son was not supporting them enough but instead supported the wife, her children, and her relatives, even if there was no evidence to support their claim. Women’s experience with their husbands’ relatives and their relatives remained one of the causes of domestic violence, as narrated by women teachers.

Although the adverse family influence was common in the narratives, it was worth noting that their husbands loved and valued their daughters-in-law in some families.

Unfortunately, in some cases, the excellent relationship with their husbands’ families brought more conflict and suffering to female teachers, as stated by some of them. FV.2 thus said:

> *His father and brothers supported me morally and were always there to protect me, but my husband became annoyed. They told him that I was not their business. After all, he was the one who brought me to their home, and therefore his family cannot be closer. He became [so] bitter that everyone, including his father and relatives, had turned against him, and supported me…”*

The above suggests that not all spouses’ relatives were hostile to married female teachers. Issues of extended families and domestic violence against female teachers were not only limited to men’s families. Female teachers cited their own family interference as one factor that leads to misunderstandings with their spouses. Some parents and families of female teachers supported their husbands to the disadvantage of female teachers.

To the teachers, their families feel that if they break up, it reflects that they had not brought them up well. Their parents want them to stick to their marriages at all costs because it was a shame when they break up. This experience points to the feminist paradigm perspective. The women say their families want them to stick even in a dysfunctional marriage because their husbands always give them a wrong impression, supporting them financially and being close to their relatives. In such situations, husbands become bolder, even more arrogant, and continually abuse them emotionally.

### Female teachers’ perspectives on the impact of domestic violence on their teaching roles

The experiences of female teachers on how domestic violence affects their teaching roles were explored. The interview interactions with female teachers and headteachers produced two themes and several sub-themes which can explain how domestic violence affects teachers’ professional and administrative roles. From these emerging themes and sub-themes, it was established that the spillover of experiences of domestic violence on female teachers gets to the workplace, affecting teaching and learning. Domestic violence leads to poor service delivery and poor interpersonal relationship with female teachers (Figure 1).

### Poor service delivery

Poor service delivery for a teacher means failure to perform in teaching and learning processes. Teachers are expected to be at school to teach all their lessons. However, domestic violence affects this as manifested by.

### Absenteeism

Female teachers’ responses indicate that domestic violence keeps them consistently away from school. Although, the actual classroom teaching needs time to be physically present to interact with students, prepare lessons, and be on call to handle any administrative duties related to teaching, this cannot be done when teachers are absent from schools. However, chronic absenteeism from duties was among the most significant effects of domestic violence on their teaching roles. Out of the 20 female teachers interviewed, (14/20, 70.0%) admitted that they absent themselves from schools after incidences of domestic violence because of physical injuries, poor emotional state, being locked up by their husbands or settling domestic issues. Out of shame and feelings of being left lonesome, they absent themselves from schools to handle and process their problems, among other issues. This was confirmed by headteachers (10/23, 43.5%) that absenteeism was common among women struggling with domestic violence.

FV.2, narrating the dilemma that sometimes she finds herself in a situation whether to go to school or stay away, lamented.

This opinion was not different from what FV.11 said,

> *It was challenging to go to school when I was in a bad mood, physical injury, and pain. The headteacher complains about my high rate of absenteeism from class. This complaint was genuine, but staying away from school after a bad day at home was better than coming to school and adding more problems. Sometimes I did not go to school because of sickness or being financially incapacitated.*

From the narratives, absenteeism from school always comes after quarrels and misunderstandings with their husbands, depriving them of sleep and lesson preparations, making them disorganized and emotionally drained. The only way out was to stay away from school while others stayed away out of fear that their spouses would follow them to school and cause a scene. Others did not go to school because their husbands had threatened to harm themselves if they “dared” leave home. In addition, physical injuries and other forms of pain made women to have excuses for staying away from schools.

> *The interaction with headteachers stressed this; one stated, *for female teachers that I suspect or have shared with me their experience of domestic violence, the level of absenteeism has been very high. Female teachers always give different excuses and tell stories to justify their absenteeism*.*

While it is true that women experiencing domestic violence were frequently absent from school, they do not come up openly to explain why they were away from work. Instead, they gave excuses. This situation meant that they did not feel good about domestic violence and felt ashamed of the situation. They reported that they stayed away because of anxiety created by the situation which affected their teaching roles even when they were in schools.

## Missing lessons

Missing lessons were among the significant effects of domestic violence on the teaching roles of female secondary school teachers. Female teachers may not be absent from school but would still miss lessons. Six (6/20, 30.0%) female teachers and five (5/23, 21.7%) headteachers said female teachers missed lessons because of late arrival to school. According to participants, they came late to school after a struggle at home, even though they knew their timetable and time to enter classrooms. They advanced several reasons for late school arrival, namely having arguments or fights just before setting off to school, being in an emotional situation that barred them from leaving home early, or sometimes they were held back either deliberately by their husbands or they were resolving conflicts. Female teachers always tried to come to school early but arrived late and often failed to enter classrooms to teach. They explained that often when they were lucky, time would be provided to compensate for the lessons missed. However, students often missed lessons even when female teachers were physically present, thus affecting syllabus coverage. Other reasons for missing lessons were lack of preparation, fatigue, not being in the mood, being emotionally drained, and disruptive behaviors from their spouses and family members.

FV.3 narrated that.

> *One day we had a fight, and my husband’s father came to school and asked the headteacher to release me from school so we could go home and handle the problem. I pleaded with him to allow me to teach and handle the domestic problem in the evening. He insisted, and the headteacher allowed me to go. I was expected to give three lessons that day but missed all. It became challenging to return to school because everyone got to know the problem. However, I got encouragement and went to work the next day. However, I had missed the lessons already, and being [a] candidate class. I should have gotten free time to compensate.*

Headteachers emphasized this problem and complained that missing lessons meant the scheme of work was not adhered to. The class lagged in such subject’s coverage, eventually making students perform poorly in exams. Such were the difficulties that female teachers faced, affecting their work at school. Being present in school, failing to perform, arriving late, and not getting to class on time were all impediments to teaching and learning as a result of domestic violence.

## Poor lesson preparations

Poor preparations were among the significant effects of domestic violence on their teaching roles. Although all participants confirmed that they knew what it took to prepare to teach; listing items such as making lesson plans from the scheme of work extracted from the teaching syllabus, making lesson notes, having learning aids and records of work, they reported that this could not be done effectively because of domestic violence. They reported that lesson preparations required time, peace of mind, and a calm environment. All these, however, could not be effectively achieved when a female teacher was experiencing domestic violence. In the interviews, fifteen (15/20, 75%) female teachers and two (2/23, 8.6%) headteachers admitted that domestic violence did not give ample time for teaching preparations. Female teachers with experience of domestic violence gave many reasons why they consistently failed to prepare their lessons well. This included stress due to many unresolved household issues, the pain inflicted by their spouses, restraint by their spouses, and the feeling of helplessness and emotional exhaustion.

They said that when they were not emotionally and psychologically prepared, they failed to prepare to teach. In addition, others got overwhelmed with family responsibilities because their husbands were either irresponsible or deliberately left all responsibilities to them. Furthermore, other participants said they failed to prepare because of the hostile home environment and disruptions of lesson preparations. Most respondents said that because of all these challenges, they gave notes or got into the classroom to occupy space and avoid reprimand from headteachers and other supervisors. Some agreed they went to a level of forging lesson plans to protect their jobs. All these can have an impact on the teaching and learning process.

FV.10 said, “*I often failed to get time to research to get the content for teaching. What I do is, I do not make lesson plans. I just walk into the class to teach*”.

For a young woman in her late 20s with minimal experience in the teaching profession, going to class without preparing well can seriously affect her teaching. Even where female teachers came up with lesson plans, they reported that they were not genuine because they just forged documents to avoid further mental torture from school administrators and, in the worst scenario losing their job, particularly those in private schools where job security was based on performance.

FV.20 says it without any remorse:

> *I always need to make lesson plans regularly and on time. To avoid reprimands from the school administrators, I always forged lesson plans by writing them when a bit settled, or after the lesson.*

Probing more on how she felt when she forged lesson plans to please the administrators and whether she could deliver effectively without adequate teaching preparations. She only laughed and responded in a very unique way.

> *Madam, you are a teacher and know what that means. Teaching is not like a soldier who does his work whether happy or not but can still fight the enemy; maybe he does it even better when he is emotional and in pain. For us teachers, we must prepare and be emotionally stable before teaching. What should I do with this man who does not give me peace of mind on the one hand and fear of losing my job?*

This finding was worrying when female teachers who were supposed to be role models and custodians of values cheated while teaching. Most women reported that their homes needed more conducive to preparing their lessons. Their husbands used this as a provocation to fight. Thus, to try and keep the peace, the women avoided working from home. One angry respondent, FV.11, in her early 30s, and married to a man of lower education level, uttered the following.

> *This man is not enlightened. Can you imagine he sometimes turns off lights when I am preparing my lessons or marking students’ scripts from home? One day he poured water on my notebook. Some friends blame me for marrying an unenlightened man. Now I agree. Back then, I thought they were irrational.*

Such utterances and attitudes can easily affect a teacher’s mental functioning, thus adversely impacting their teaching. FV.3, narrating her experience during the interview, said,

> *One day in the evening, after supper, I was making a lesson scheme of work to meet the deadline in school. He called me, and I went and knelt near him. He just looked at me and kept quiet. I immediately sensed danger. He asked me to get out of his sight and leave him alone. I left and went to pick [up] my books and left for the bedroom. He followed me, started twisting my arm, and beat me up. In such a condition, can any woman be fully prepared to teach?*

Further probing into this led to two female teachers in their 40s giving similar reasons saying their husbands think when they are preparing lessons in the evening, they do it to annoy, deny them attention, and punish them. That is why they disrupt lesson preparations. That the husbands say they should finish schoolwork at school.

The researcher was interested in these two participants and asked whether the allegations from their husbands were true.

One of them, FV.19, answered without hesitation, saying, “*I will do anything to avoid getting intimate with him. I am tired, and God knows! “Staying up late to make lesson plans is a good excuse*.” However, this breeds more conflict. Headteachers re-emphasized the above point. One stated:

> *Teaching lessons without a marking scheme of work, lesson plans and without lesson notes, but instead reading information directly from pamphlets to learners affects content delivery as it can always be reflected in poor performance in subjects of such teacher at UNEB ordinary level examinations. That explains why female teachers fail to prepare to teach when they are not emotionally and psychologically settled. Others failed to prepare because their spouses completely neglected their roles, and the entire burden lay with the women. In such a situation, their work as teachers was affected.*

Although up to 75% of the women interviewed recognized that domestic violence had a bearing on lesson preparations, not many headteachers took this seriously. This issue probably emerged from the lack of supervision, where headteachers were not closely following what took place in the classrooms.

## Low self-esteem

Low self-esteem was among the significant effects of domestic violence on female teachers’ roles. A teacher requires confidence to enable her to perform well, yet consistent abuses and fights made them develop low self-esteem with diminished productivity. Up to nine (9/20, 45.0%) female teachers interviewed mentioned that spousal conflicts made them to develop low self-esteem. They questioned their worthiness and doubted their ability to perform anything good. Accordingly, they developed poor self-images because of consistent abuses, negative comments, ridicules, and belittling from their spouses. The low self-esteem also came from the lack of a peaceful mind, emerging regrets, and shame. Some older women, however, said that with time they reclaimed their self-image and confidence after spiritual support, reassurance from friends, and counseling, although it became counterproductive because they become sensitive to any comments or when they sensed someone wanting to interfere with their freedom. This experience made them develop poor interpersonal relations with others, thus affecting their teaching roles.

Three headteachers (3/23,13.0%) also observed this as accurate and agreed with the observations. The difference between headteachers and female teachers who share the same may have been because most headteachers did not note low self-esteem on female teachers who remained strong despite the many challenges. In contrast, higher percentages of female teachers than headteachers reported their perceptions through their stories.

Some confirmed this. FV.19, a teacher in her 40s, had this to say:

> *I have developed a poor self-image. I would ask him if I were [if I was] so ugly. Nevertheless, his response would annoy me so much. One day in my rage, I asked him: “Is there any value at all that you see in me?” He responded: “What do you think? Would I be running away from you if you still had anything for me to admire?” I felt so bad. I wanted to die. I repeatedly asked myself if I was that useless and had any value as a teacher.*

Those who said they found solutions and regained self-esteem said they performed their professional roles well. FV.13, who had a harrowing experience, having lived with two men, and separated, said:

> *I had low self-esteem because of what I went through. I thank God that I became a trained counselor, and since then, I have also become helpful to myself and others. I do not know where it would have ended. Maybe he would have killed me by now. Before joining the course, I became abusive and never trusted him. I would begin to fight anytime, and most times when he was away, I would be planning only how to hurt him so that he would feel pain as well, and this affected me as a teacher— however, this increased violence and hostility in the house. I only stopped this when I became saved, and that, of course, came when I got support from the lecturer and my course-mates during the course, and then I became saved.*

It was, however, difficult to establish how many such women took things positively. Further narratives were from FV.14:

> *Teaching requires a very calm and peaceful mind. When I started teaching, I was very hardworking. I would reach school on time, prepare all my lessons, and teach all my lessons. However, what I have gone through has changed my work pattern, and sometimes I nurse regrets and shame.*

## Mental health problems

To prepare lessons, deliver content, interact with students, and interact with learners’ work, a teacher must be mentally sound and motivated, yet domestic violence affects the mental functioning of a teacher. In this study, several manifestations from the participants point to some problems that barred them from performing their teaching roles effectively. Some of these were low motivation (4//20, 5.0%), loss of concentration (9/20, 45.0%), sleep problems (5/20, 25.0%), fatigue (7/20, 35.0%), isolation (11/20, 55.0%), among others. The mental states were due to the hostile and toxic household environment. They reported that being at home never gave female teachers the peace of mind.

Another reason that affected their mental concerns was knowing that innocent children were suffering, neglected, and hurt, yet failing to see how they could escape the situation. These were stated by the female teachers interviewed. FV.15, said,

> *At the peak of our conflict, I would travel from school and go home, even if I planned to spend the whole weekend at home, but the toxic home environment would always make me return to my workplace. Of course, much as I was present in school, I would remain hurt and mentally unsettle. This situation affects my work substantially. I remember one day I came home and found him in a very aggressive mode. The moment I greeted him, he started fighting and that became too much for me. I immediately left home for school. My children cried and pleaded with me not to leave. I could not bear this anymore. When I reached the school, I stayed awake and wept throughout the night. In days and weeks to follow, each time I was making lesson plans or sometimes in the classroom teaching, I would see my children vividly in my mind crying and asking me not to leave them. I love my children; I love my work too, but I knew living with this man would make me lose my job and affect me physically and emotionally.*

## Poor interpersonal relationships

Poor interpersonal relationships emerged as another theme that affect female teachers’ professional and administrative roles. A teacher’s work involves interaction with many groups of people, remarkably with colleagues, staff, and students. For a teacher to perform her work effectively, she must have excellent interpersonal relationships with those she interacts with, yet domestic violence adversely impacts this, thus affecting their teaching roles (Figure 2).

## Bad attitudes

Female teachers reported that because of their household experience of domestic violence, they often develop bad attitudes towards their husbands and deflect them onto others. In schools, students and other staff become victims of such attitudes. Female teachers reported that when unhappy or stressed, they displaced their hostilities on students by becoming very harsh and rude. Sometimes, they get irritated by simple things, become aggressive to the extent of beating or chasing students out of class. To female teachers, this often happened without intent, but usually on circumstances beyond their control. Nonetheless, one participant, in her late 30s, said she had developed a hatred for men, and sometimes the boys in her class became victims of her bad attitudes. This experience affected their relationships with other teachers and students. It was noted that students begin to fear, avoiding and hating the subjects they taught, or avoiding their classes altogether.

On the other hand, female teachers admitted that often they realized how aggressive they had been and regret punishing students innocently, which further affected their productivity and peace of mind.

## Lack of cooperation

Most female teachers reported that domestic violence negative affected their relationships with their workmates. When asked to explain this, they said that the lack of cooperation with other teachers were because of their unusual behaviors, where, in most cases, they were easily irritated, and fellow workers would avoid and stay away from them. They agreed that they usually failed to work as a team on tasks assigned because, they were not available or were not in the mood to work. In this case, the other teachers would feel overloaded because of the gaps they created.

Most headteachers confirmed that female teachers with experience of domestic violence had abysmal interpersonal relations with the rest of the teachers because they were uncooperative and liked misplacing their hostilities with other innocent teachers. Besides, they also fail to guide learners, and, as such, this affects their teaching roles. These were some of the responses by participants who showed unusual attitudes among the reasons that negatively affected their teaching roles.

FV.19 told the researcher, *“I must confess that I always become so aggressive on students for no justifiable reason, and when I reflect later, I realize that my reaction to them was extreme. I only feel too ashamed to go back to apologize to them”*.

FV.11 reported that *“…many times, when I go to school after a disagreement with my husband, I find myself very harsh and rude to students. Sometimes I become arrogant, especially when students are asking difficult questions”*.

These revelations reflect the relationship between female teachers and their students. It was unusual that teachers who must be close to their students, to guide and support them find themselves not supportive of the students. Such students begin to avoid the teacher, their lessons, and later respond by hating the teacher, and soon the rest of the students joined in too. In such circumstances, teaching and learning usually do not go on well as expected of student-teacher relationship.

Poor interpersonal relations occurred when teachers affected by domestic violence failed to cooperate with other teachers. FV.5, speaking about interpersonal relations with other co-workers, said:

> *As a teacher, we must always work as a team in our department. Geography is one such department that requires teamwork. Most often, I want to sit alone and do my things alone. I do not know why. I used to be a team player and loved to socialize. Unfortunately, this is no more.*

Getting isolated and failing to work as a team means such a teacher could not be a resource to their colleagues in the department and could not even get support from others. The narrative in the interview reveals that with time tension develops with the other teachers, and gossip sets in, causing the environment unbearable for teachers who are already suffering from domestic violence. Students in the class become the loser in these circumstances.

FV.11 expressed how domestic violence kept her isolated from other co-workers at school. She said, *“I need help with relating even with my colleagues. I do not greet or stay in the staff room with them, which stresses me. I always feel that everyone is laughing at me. Now they have also developed [an] attitude on me. When I feel like being with them or wanting their support, they are often unwilling to relate quickly. I do not blame them. It is my fault*.

Concerning the above, headteachers stated that teachers fail to guide learners due to domestic violence. One stated, *“As teachers, we are supposed to guide learners. Teachers in domestic violent relationships are not free with learners, and very few learners come to them for support outside the class hours”*. This finding means that women undergoing domestic problems cannot sustain a good relationship with learners.

## Discussion

This study established that the spillover of experiences of domestic violence on female teachers got to the workplace and affected teachings and learning (Figure 1, Figure 2, Figure 3). This finding agrees with a study conducted by Melsa (2014), where 92% of the women agreed that domestic violence considerably impact their work.^25^

These findings reveal that all teachers were aware of their teaching roles. Teachers play multiple and complex roles, which are crucial and require commitment and focus if teaching and learning are to be effective on learners. However, the study revealed that domestic violence interfered with the smooth implementation of those roles (Figure 2). A teacher who performs her work well must be stable and free from domestic violence, particularly when we consider the vital roles of teachers in facilitating learning of students. The study revealed several factors hindering female teachers’ performances because of their experiences of domestic violence (Figure1, Figure 2 and Figure 3).

Poor service delivery was observed among female teachers in abusive relationships as evidenced by persistent absenteeism, missing lessons, and poor lesson preparations before classes (Figure 1).

Further the contributing factors to poor service delivery were low self-esteem that female teachers developed or perceived because of domestic violence and mental instability that barred them from performing their roles effectively. This was because domestic violence causes them to lose concentration, sleep, increase fatigue, and isolation. In this study, the effects of domestic violence mentioned above were identified as contributors to poor service delivery and thus affected female teachers’ performances with lived experiences of domestic violence (Figure 1). These findings were consistent with earlier studies on the impact of domestic violence on productivity of working women.^27,28,29^

This study revealed that violence inflicted by husbands of female teachers affect their capacity to work. Female teachers said they reported late, were persistently absent or left school early, more specifically during challenging times, and some were done as a strategy to avoid violence at home. Consistent with our study, persistent absenteeism from work by female teachers with lived experiences of domestic violence was reported in other studies.^31,32,36^ Other studies also revealed that employers were disadvantaged as it was difficult to maintain productive operations when employees were consistently absent, tardy, or displayed low productivity levels.^39^ Therefore, it does not matter what kind of institution, but persistent absenteeism affects workplace performance, as evidenced by this study in secondary schools in Uganda. This study was consistent with other studies on educational institutions focusing on the absenteeism of teachers and implications for students’ achievements, where studies reported that the relationship between teachers’ absence and students’ achievement indicate that teachers’ absence had a statistically significant negative relationship with students standardized mean academic achievements.^22,40^

This finding implies that female teachers who absent themselves from school because of domestic violence adversely affect students’ learning achievements because they fail to teach and cover the syllabus on time; they were inconsistent in attendance and failed to make themselves available in schools to guide students.

Although students’ performance cannot be limited to just one variable of persistent absenteeism of teachers, we could not ignore it for this study because even after being away from school, female teachers still returned to school in an unstable mind and without adequately preparing their lessons. This situation was because they were recovering from past incidents of domestic violence or in anticipation of the next move of their spouses hurting them and how they could avoid them. This experience can well be understood in the motivational theory advanced by Maslow’s hierarchy of needs, where female teachers were desirous of safety and could not get it from their spouses. So, this would not motivate them to perform well in schools.^41^

This study also revealed that teachers’ presence at school did not necessarily translate into effective teachings and learnings. Female teachers came to school unprepared, needing more concentration and simply being present to protect their jobs, but this did not translate into good performance. The lack of concentration was in line with other studies conducted among nurses, where ½ of survivors felt less able to concentrate on their work and could not work to the best of their ability, and many took sick leaves.^34^ On the other hand, administrators were more concerned with their physical presence in schools, as revealed by this study. These findings were in contrast with studies that brought in key roles of school supervision by administrators in both pedagogical and administrative duties of teachers to ensure good performance.^26^ Headteachers can not rate the performance of workers by their physical presence only; but must closely monitor their level of preparation, delivery, assessments, and relationships with students, and once this is not done consistently, teachers’ performances will remain unchecked.

Several studies have pointed out that the behaviors of spouses have overwhelming effects on the work performance of their spouses. Victims feel unwell, tired, and distracted from duties.^27,35^

Similarly, a report produced by Australia National Domestic Violence and Workplace Survey (2011) showed that ½ of respondents from 3600 populations who faced domestic violence admitted that violence affected their capacity to work.^42^

Consistent with this current study’s findings, female teachers agreed that their capacity to work was affected because of lived experience of domestic violence they experienced making them lose self-esteem and feeling that they did not have any worth. This feeling of worthlessness made them lose confidence even with students. They often felt isolated and did not correctly interact with co-workers.

According to Maslow’s Hierarchy of Needs, humans desire esteem needs for self-esteem and self-respect.^41^ They need to attain freedom and independence that give them confidence as they continue to do their work. They desire respect from others, recognition, and appreciation for what and who they are.^37^ When people feel respected, they develop a sense of self-worth, enabling them to perform well in their workplaces.

Furthermore, Domestic violence often adversely affect female teachers who feel their spouses do not respect them and therefore feel worthless. They lose self-confidence that is critical to a teacher’s professional roles, which affects their productivity unless a good coping measure is provided.

This study contrasts studies conducted by Rayner-Thomas (2013)^4^ and Swanberg et al. (2005),^36^ where they noted that gender organizations assumed that employers were unable and unwilling to accept that, what happens in an employee’s home does not invariably come to workplaces and affect their performance.^4,36^ The same view was shared by Meinert (2017), who showed that 71% participants in his study showed that domestic violence was not a problem at their respective companies.^14^ However, most participants from our study agreed that domestic violence affect their teaching performance in schools.

A few participants said the violence inflicted on them from home did not affect their teaching and administrative roles. This finding may be insignificant since these responses came only in one aspect of games and sports, which could be observed as a therapy to one female teacher, who, after facing domestic violence from her husband, found pleasure in doing sporting activities with students.

In all, domestic violence affects the work of female secondary schoolteachers. Any contrasting views to this, in our opinion, points to views that continue to make women educationalists suffer during their work with a resulting poor performance.

This study showed that all female teachers were aware of the harmful effects of domestic violence but were unwilling to make it hinder their professional performances. Crucially, where organizations and institutions feel that domestic violence cannot have a spillover in workplaces like schools, they fail to address the challenges faced by female schoolteachers.

In secondary school settings, when a teacher fails to discuss matters about the subject one teaches with colleagues or department members and where one is not free with students, teaching and learning become a challenge. Fatigue and lack of sleep were disruptive to lesson preparations and effective delivery of teachings and learning. Therefore, where adequate preparation was not done for teaching, where a teacher came late to class or missed lessons, curriculum delivery was affected, and cumulatively, it affected the performance of students and the school in national examinations.

We, the authors, argue that domestic violence must be addressed so that the spillover effects do not significantly affect the performance of female schoolteachers and students they teach. Other studies were consistent with ours in that domestic violence can significantly impact employees’ productivity and work, which becomes prominent when female teachers are experiencing domestic violence.^38,43,44^ Experience from elsewhere in the African continent show that domestic violence affects female productivity and ultimately their overall performance and the organization at large.^38^ It is reported that productivity of victims experiencing domestic violence is reduced due to sapping of their energy, undermining their confidence, and compromising their health.^43^ In addition, domestic violence leads to persistent absenteeism, loss of work time, high labour turnover, and low productivity.^44^

Inconsistent with our findings, a study reported that women who no longer experience intimate partners’ violence exerted less or no work distraction.^14^ This finding contrasts with ours but highlights the impact of lived experience of domestic violence on females who left relationships but were still preoccupied with memories of negative expressions made by their partners, mistreatments of the past, reminders of challenges of managing children, and regrets.

Consistent with our study, another reported that physical aggression predicted higher levels of withdrawal both at and from work, with psychological aggression predicting additional variance in partial absenteeism over and above the effects of physical aggression.^29^ Last, consistent with our findings, two studies show that intimate partner aggression reflects behaviors that are intended to boost and maintain male superiority while enhancing female dependence.^44,45^ It becomes more critical in such a situation that women are engaged in behaviors that maintain their economic independence for example arriving at work on time and remaining productive.

What appears to determine these findings among female schoolteachers was the enabling environment or lack of it, the severity and duration of the abuse, among other challenges faced by female schoolteachers in this study population. We, the authors, propose that these are areas for further studies.

## Limitations of this study

Although this study has provided key findings and information about perceptions and lived experiences of domestic violence among female secondary schoolteachers and its effects on the teaching roles in secondary schools, there are limitations to be considered.

Considering the number of study participants, it may be inadequate to indicate workplace implications stemming from domestic violence inflicted on employees and to draw general conclusions from these findings. However, the study highlighted the need for employers to find strategies to address spillovers of domestic violence in schools. Besides, the generalisability is still valued because participants were teachers who received training from similar institutions, and the issue of domestic violence is a national issue; therefore, the findings can be generalized to other areas too.

Second, snowball as a sampling method has several challenges, which could affect the research findings. There were concerns that those teachers with a smaller network may be underrepresented, as they were less likely to be referred to for interviewers, yet their perspectives could be of interest in this study.

## Strengths of this study

This study had many strengths.

First, regular contacts, being close to participants, and clearly explaining the purpose of the study helped us to address some of the shortcomings of the study sampling method. The research team gained confidence, trust and referred other participants for the study thus reaching all potential victims of domestic violence in the area.

Second, although the number of participants was small, in-depth interviews allowed the research team to gain valuable information until theoretical saturation was reached. Besides, the in-depth interview provided the opportunity to get richer data and in-depth insights by exploring women’s experiences of domestic violence and how it affects them.

Third, the data was based solely on self-reporting by participants; female teachers, and headteachers. Triangulating information with administrators was important in enriching the information obtained and crosschecking the authenticity of some data from the victims. Fourth, interviews with other family members or co-workers of the victims (female secondary school teachers) gave the study more understandings of the effects of domestic violence on their professional and administrative roles.

Fifth, the sensitivity of this topic made the researcher conduct interviews, code findings, and analyze the facts without exposing personal information to others. This approach could be a potential source of bias. In order to address any possible biases, several strategies were applied to limit bias reporting. These included audit trail, peer debriefing, flexibility and triangulation of information obtained.

Sixth, this data was collected during the peak of the COVID-19 pandemic; a time that was so stressful, and people lived in fear and with financial difficulties. Most teachers from private schools were not receiving salaries at the time. To overcome this, we gave hope and lifted their spirits by candid conversations on domestic violence, and by doing so, we motivated them to narrate their in-depth stories on domestic violence. The advantage it gave us was the long available time for engagements with schoolteachers since schools were closed and the long hours for conversations were available.

## Generalizability

Although this study handled a small number of participants and only in secondary schools in Kitgum District, findings are cautiously generalizable to a broader population of female secondary school teachers in Uganda who are victims of domestic violence. This finding includes generalization through context similarity of the African culture that remains patriarchal with men having control over women and that such powers are exerted through force. Therefore, issues of domestic violence may be similar throughout the country. In addition, the education management system applies to all schools in Uganda (Private or Government Schools). The Acts, policies, and regulations governing the management of secondary schools are the same and teachers and administrators are all trained in the same institutions using the same curriculum and are, therefore, most likely to behave similarly. In addition, Uganda’s legal system and other laws are the same and therefore are bound to apply in the same way, whether in strengths or weaknesses. Furthermore, this study is underpinned by a feminist theory, a universally accepted theory used by researchers studying domestic violence. Because of this, the findings may be applied to a broader context.

## Conclusion

This exploratory study brings awareness to lived experiences and the effects of domestic violence on female secondary school teachers in Uganda. Domestic violence affects their teaching and administrative roles with resulting poor service delivery and poor interpersonal relationships. Providing a supportive work environment is crucial for female teachers experiencing domestic violence to enable them to perform their professional and administrative roles. Different players, including school managers, the Ministry of Education and Sports, feminist organizations, and relevant government offices, can intervene by establishing workplace policies and relevant processes to support female teachers who experience domestic violence.

## Declarations

### Ethics approval and consent to participate

Uganda Christian University (UCU) and Uganda National Council for Science and Technology (UNCS&T) approved our study. We obtained written informed consent from all participants. In addition, the study was conducted following all relevant institutional guidelines and regulations.

## Consent to publish

Participants provided written informed consent.

## Availability of data and material

All datasets supporting this article’s conclusion are within this paper and are accessible by a reasonable request to the corresponding author.

## Authors’ contributions

GAO, MOK, and WE designed this study— GAO, MOK, and WE supervised data management. GAO, MOK, DLK, and WE analyzed and interpreted the data. GAO, MOK, DLK, and WE wrote and revised the manuscript. All Authors approved the manuscript.

## Authors’ Information

Gladys Ayot Oyat (GAO) is a Gulu University Council member and a former headteacher of YY Okot Memorial College, Kitgum, Uganda; Dr. Mary Ocheng Kagoire (MOK) is a consultant at Uganda Management Institute (UMI), Kampala, Uganda; Dr. Wilson Eduan (WE) is a senior Lecture at Uganda Christian University, Mukono, Uganda; Prof. David Lagoro Kitara (DLK) is a Takemi fellow of Harvard University and a Professor at Gulu University, Faculty of Medicine, Department of Surgery, Gulu City, Uganda.

## Abbreviations

FV: Female Victim
IPC: infection, prevention, and control
IRB: Institutional Review Board
SOPS: Standard Operating procedures
UCU: Uganda Christian University
WHO: World Health Organization
COVID-19: Coronavirus disease-19

## Acknowledgment

We thank the assistance of the family and the team that supported the review and typesetting of this work.

## References

1. Shorey S, Chua CMS, Chan V, Chee CYI. Women living with domestic violence: Ecological framework-guided qualitative systematic review. Aggression and Violent Behavior. 2023;71:2023,1e01835.

2. Swallow J. An exploratory study of women’s experiences regarding the interplay between domestic violence and abuse and sports events (Doctoral dissertation). University of Chester, United Kingdom. 2017.

3. Garcia-Moreno C, Jansen A F MH, Ellsberg M, Heise L & Watts C. WHO Multi-country study on women’s health and domestic violence against women. World Health Organisation. 2005.

4. Rayner-Thomas MM. The Impacts of Domestic Violence on Workers and the Workplace. University of Auckland [Preprint]. 2013. Available at: https://www.ituc-csi.org/IMG/pdf/the_impacts_of_domestic_violence_on_workers_and_the_workplace_-_margaret_rayner-thomas.pdf.

5. LAWS OF MALAYSIA Act 795 access to biological resources and benefit sharing act 2017. Percetakan Nasional Malaysia Berhad. 2017. https://chrome-extension://efaidnbmnnnibpcajpcglclefindmkaj/https://faolex.fao.org/docs/pdf/mal176890.pdf

6. Nyangoma P. An analysis of domestic violence problems in Uganda. Department of Journalism and Communication-Makerere University. 2012. https://jocom.mak.ac.ug/news/analysis-domestic-violence-problem-uganda.

7. Ararat M, Alkan S, Budan P, Bayazıt M, and Yüksel A. Domestic violence against white-collar working women in Turkey: a call for business action. Sabancı University. 2014. ID:10.5900/SU_SOM_WP.2014.25972. https://research.sabanciuniv.edu/25972/1/BADV_Report.pdf Available at: 10.5900/su_som_wp.2014.25972.

8. Koenig MA, Lutal T, Zhao F, Nalugoda F, Wabwire-Mangen F, Kiwanuka N, et al. Domestic violence in rural Uganda: Evidence from a community-based study. Bull World Health Organisation.2003;81(1):53–60.

9. El-Zanaty F, Hussein EM, Shawky G, Way A and Kishor S. Egypt Demographic and Health Survey. National Population Council and Macro International.1995.

10. United Nations Population Fund (UNFPA). Evaluation of UNFPA support and the prevention, response to the elimination of gender base violence and harm practices. 2018. https://www.unfpa.org/sites/default/files/adminresource/GBV_Report_FINAL_29_Nov.pdf.

11. Ministry of Gender, Labour and Social Development Ministry of Gender, Labour and Social Development, National Action Plan on elimination of Gender-Based Violence in Uganda (2016-2020) ISBN: 978-9970-507-18-4.

12. Stephen Ssenkaaba. Uganda: Violence against women unabated despite laws and policies. 2018. https://www.un.org/africarenewal/news/uganda-violence-against-women-unabated-despite-laws-and-policies

13. Campbell JC, Anderson JC, McFadgion A, Gill J, Zink E, Patch M, Callwood G, Campbell D. The Effects of Intimate Partner Violence and Probable Traumatic Brain Injury on Central Nervous System Symptoms. J Women Health (Larchmt). 2018;27(6):761–767. doi: 10.1089/jwh.2016.6311.

14. Black E, Worth H, Clarke S. Prevalence, and correlates of intimate partner violence against women in conflict affected northern Uganda: a cross-sectional study. Confl Health. 2019;13:35.

15. Uganda Demographic Health Survey (UDHS). Uganda Bureau of Statistics Kampala, Uganda, MEASURE DHS ICF International Calverton, Maryland, USA. 2012. http://chrome-extension://efaidnbmnnnibpcajpcglclefindmkaj/https://dhsprogram.com/pubs/pdf/fr264/fr264.pdf

16. Uganda Bureau of Statistics (UBOS). Uganda demographic and health survey. 2011.

17. Uganda Demographic Health Survey (UDHS). Uganda Bureau of Statistics Kampala, Uganda, MEASURE DHS ICF International Calverton, Maryland, USA. 2016.

18. Kitara DL, Odongkara BM, Anywar DA, Atim P, Amone C & Komakech D. Domestic violence in Gulu-Northern Uganda. East and Centr Afr J Surg.2012;17(1):29–36.

19. Riessman CK. Narrative analysis. Sage Publications. 2005.

20. Naderifar M. Snowball sampling: A purposeful method of sampling in qualitative research Strides in Development of Medical Education. 2017.

21. Unicef. Education for every child, an education. 2020. https://www.unicef.org/uganda/what-we-do/education

22. Bold C. Using Narrative in Research. Sage Publications. 2011.

23. Creswell JW, Poth CN. Qualitative Inquiry & Research Design: Choosing among 5 approaches. Fourth Edition. Sage Publications, Inc. 2017.

24. Brink HIL. Validity and Reliability in Qualitative Research. Curationis.1993;16(2):a1396. DOI: 10.4102/curationis.v16i2.1396.

25. Stewart DE, MacMillan H, Kimber M. Recognizing and Responding to Intimate Partner Violence: An Update. The Canadian Journal of Psychiatry. 2021;66(1):71–106. doi:10.1177/0706743720939676.

26. Sule Mary Anike, Ameh Eyiene, Egbai Mercy E. Instructional Supervisory Practices and Teachers’ Role Effectiveness in Public Secondary Schools in Calabar South Local Government Area of Cross River State. Nigeria J Education and Practice. 2015;6(23). www.iiste.org ISSN 2222-1735 (Paper) ISSN 2222-288X.

27. Swanberg JE, Macke C, & Logan TK. Intimate Partner Violence, Women, and Work: Coping on the Job. Violence and Victims. 2006;21(5):561–578.

28. Alaniz R & De Los Santos E. Domestic violence: It can happen to professional women including educators. International Journal of Education and Social Science.2015;2(12):1–10.

29. LeBlanc MM, Barling J & Turner N. Intimate partner aggression and women’s work outcomes. Journal of Occupational Health Psychology.2014;19(4):399–412.

30. Swanberg JE, Logan TK & Macke C. Intimate partner violence, employment, and the workplace: Consequences and future directions. Trauma, Violence & Abuse. 2005;6(4):286–312.

31. Scott KL, Lim DB, Kelly T, Holmes M, MacQuarrie BJ, Wathen CN, et al. Domestic violence at the workplace: Investigating the impact of domestic violence perpetration workers and workplaces. Toronto, ON: University of Toronto. 2017.

32. Reeves C and O’Leary-Kelly AM. The effects and costs of intimate partner violence for work organizations. Journal of Interpersonal Violence. 2007;22(3):327–344.

33. Abrahams, H. Supporting women after domestic violence, Trauma and Recovery. Jessica Kingsley Publisher, 1^st^ edition. London NI 98, UK. 2007.

34. Gupta M, Shaheen M and Reddy PC. Impact of psychological capital on organizational citizenship behavior. Journal of Management Development. 2017;36(7):973–983.

35. McFerran L. Domestic violence workplace rights and entitlement project: National Domestic Violence and the Workplace Survey: Australian Domestic and Family Violence Clearing House. 2011. http://dvatworknet.org/sites/dvatworknet.org/files/Australia_survey_report_nov2011.pdf

36. Barnett OW. Why Battered Women Do Not Leave, Part 2: External Inhibiting Factors-Social Support and Internal Inhibiting Factors. Trauma, Violence, & Abuse.2001;2(1):3–35. 10.1177/1524838001002001001.

37. Margaret Michelle Rayner-Thomas. The Impacts of Domestic Violence on Workers and the Workplace. A thesis submitted in partial fulfilment of the requirements for the degree of Master of Public Health (MPH), The University of Auckland, 2013. https://chrome-extension://efaidnbmnnnibpcajpcglclefindmkaj/https://zonta.org.nz/wp-content/uploads/2015/10/The-Impacts-of-Domestic-Violence-on-Workers-and-the-Workplace-Margaret-Rayner-Thomas.pdf.

38. E.E. Oni-Ojo, Anthonia Adeniji, Adewale Omotayo Osibanjo, TP Heirsmac.

39. Kelly JTD, Colantuoni E, Robinson C. From the battlefield to the bedroom: a multilevel analysis of the links between political conflict and intimate partner violence in Liberia. BMJ Glob Health 2018;3:e000668. doi:10.1136/bmjgh-2017-000668.

40. Muhammad Ameeq, Muhammad Muneeb Hassan, Laraib Fatima. Impact of Teacher Absenteeism on Student Achievement: A Case of South Punjab District Muzaffargarh, Pakistan. Journal of Education and Practice. 2018;9(16) – www.iiste.org ISSN 2222-1735 (Paper) ISSN 2222-288X.

41. Saul Mcleod and Saul Mcleod. Maslow’s Hierarchy of Needs. 2023. https://www.simplypsychology.org/maslow.html#:~:text=From%20the%20bottom%20of%20the,can%20attend%20to%20higher%20needs.

42. Australian Bureau of Statistics (ABS). Australia National Domestic Violence and Workplace Survey in 2011. Directory of Family and Domestic Violence Statistics. 2013. https://www.abs.gov.au/ausstats/abs@.nsf/Lookup/4533.0Main+Features472013.

43. Ajala EM. Domestic Violence and the Workplace: Improving Workers’ Productivity. A Journal of Contemporary Research. 2013;10(1):129–144, 2013 ISSN:1813-2227.

44. Emmanuel Majekodunmi Ajala. Impact of Domestic Violence on the Workplace and Workers’ Productivity in Selected Industries in Nigeria. Anthropologist. 2008;10(4):257–264. DOI:10.1080/09720073.2008.11891059.

45. Tolman RM & Wang HC. Domestic violence and women’s employment: Fixed effects models of three waves of women’s employment study data. Am J Community Psychology. 2005;36:147–158. doi:10.1007/s10464-005-6239-0.

46. Yodanis CL. Gender inequality, violence against women, and fear: A cross-national test of the feminist theory of violence against women. J Interpersonal Violence. 2004;19:655–675. doi:10.1177/0886260504263868

